# Clinical characteristics of critically ill patients with COVID-19

**DOI:** 10.1101/2020.12.09.20246413

**Authors:** Indalecio Carboni Bisso, Iván Huespe, Carolina Lockhart, Agustín Massó, Julieta González Anaya, Micaela Hornos, Romina Famiglietti, Marcelo Di Grazia, Pablo Coria, Eduardo San Román, Marcos Las Heras

## Abstract

**Objective:** Describe the clinical and respiratory characteristics of critical patients with coronavirus disease 2019 (COVID-19).

**Design:** Observational and retrospective study over 6 months.

**Setting:** Intensive care unit (ICU) of a high complexity hospital in Buenos Aires, Argentina.

**Patients:** Patients older than 18 years with laboratory-confirmed COVID-19 by reverse transcriptase-polymerase chain reaction (RT-PCR) for SARS-CoV-2 were included in the study.

**Variables of interest:** Demographic characteristics such as sex and age, comorbidities, laboratory results, imaging results, ventilatory mechanics data, complications, and mortality were recorded.

**Results:** A total of 168 critically ill patients with COVID-19 were included. 66% were men with a median age of 65 years (58-75. 79.7% had at least one comorbidity. The most frequent comorbidity was arterial hypertension, affecting 52.4% of the patients. 67.9 % required invasive mechanical ventilation (MV), and no patient was treated with non-invasive ventilation. Most of the patients in MV (73.7%) required neuromuscular blockade due to severe hypoxemia. 36% of patients were ventilated in the prone position. The length of stay in the ICU was 13 days (6-24) and the mortality in the ICU was 25%.

**Conclusions:** In this study of critical patients infected by SARS-CoV-2 in a high-complexity hospital, the majority were comorbid elderly men, a large percentage required invasive mechanical ventilation, and ICU mortality was 25%.

## INTRODUCTION

In December 2019, a new coronavirus was identified by the Chinese Center for Disease Control and Prevention. On March 11th of 2020, the World Health Organization (WHO) declared the severe acute respiratory syndrome coronavirus 2 (SARS-CoV-2) outbreak as a pandemic due to the constantly increasing number of cases outside China^1^. Up to date, SARS-CoV-2 affected more than 32 million people over the world and caused 1 million deaths.

On March 3rd of 2020, 64 days after the first case was reported in China, a case in Argentina was confirmed. Since then the number of cases gently ascended^2^; up to September 30th of 2020, a total of 736.609 had tested positive for the new SARS-CoV-2, with 139.419 active cases and 3.792 (2.7%) admitted to intensive care unit (ICU)^3^.

Nevertheless, local information about the incidence and clinical characteristics of critically ill patients diagnosed with COVID-19 is still limited^4^. In this context, knowledge of the baseline characteristics and outcomes of critically ill patients is crucial for health and government officials engaged in planning efforts to address local outbreaks. This case series describes clinical characteristics, image findings and respiratory mechanics of COVID-19 patients admitted to ICU in a High Complexity Hospital in the city of Buenos Aires.

## METHOD

### Study design and participants

For this retrospective single-center study, ICU patients were recruited between March 15th of 2020 and September 15th of 2020. Data were obtained from medical records of adult patients (18 years of age or older) with laboratory-confirmed COVID-19, hospitalized in the ICU in a High Complexity Hospital in the city of Buenos Aires. Patients with adequacy of therapeutic effort at ICU admission were excluded.

According to WHO guidance^5^, laboratory confirmation for SARS-CoV-2 was defined as a positive result of real-time reverse transcriptase-polymerase chain reaction (RT-PCR) assay of nasopharyngeal swabs. Only laboratory-confirmed cases were included in the analysis and the study was approved by the Hospital Ethics Committee in March 2019.

### Procedure

Epidemiological, demographic, clinical, laboratory, respiratory support, and outcome data were obtained. Radiologic assessment of chest x-rays and all laboratory testing was performed according to treating physician criteria. The presence of a radiologic abnormality was determined and the Radiographic Assessment of Lung Edema (RALE) score^6^ was calculated and reviewed by medical imaging specialists. Laboratory assessment consisted of a complete blood count, a blood chemical analysis, coagulation testing, assessment of liver and renal function, and measure of electrolytes, C-reactive protein (CRP), procalcitonin, pro b-type natriuretic peptide (pro-BNP), lactate dehydrogenase, high-sensitivity cardiac troponin (hs-cTnT), d-dimer, ferritin and interleukin-6 (IL-6).

The number of patients who had died or had been discharged, and those that stayed in the ICU until September 30th of 2020 was recorded. Additionally, ICU length of stay was determined.

### Statistical analysis

No statistical sample size calculation was performed in advance and the sample size was equal to the number of patients treated during the study period. Continuous variables were expressed as medians and interquartile ranges or simple ranges, as appropriate. Categorical variables were summarized as counts and percentages.

No imputation was made for missing data. Mann-Whitney rank-sum test was used to compare nonparametric continuous variables. χ2 or Fisher exact test was used for categorical variables as appropriate. All statistical tests were 2-tailed, and statistical significance was defined as P < .05. The analysis has not been adjusted for multiple comparisons, and given the possibility of a type I error, the findings should be interpreted as exploratory and descriptive. All the analyses were performed with the use of R Software, version 3.6.2 (R Foundation for Statistical Computing).

## RESULTS

### Demographic and Clinical Characteristics

Between March 15th of 2020 and September 15th of 2020, a total of 259 patients with suspected COVID-19 were referred to the ICU in a high complexity hospital in the city of Buenos Aires. A total of 87 patients with negative test results for SARS-CoV-2 were not included in the study. Additionally, 4 patients with laboratory-confirmed COVID-19 but adequacy of therapeutic effort at ICU admission were excluded too. Thus, data from 168 critically ill patients with laboratory-confirmed COVID-19 was analyzed.

Overall, 66% (111) patients were male with a median age of 67(58-75) years old. Eighty percent (134) of patients presented at least one comorbidity. Hypertension was the most common comorbidity, affecting 52,4% (88) of patients, followed by obesity in 41,6% (70) patients. 6.5% (11) of patients admitted were health-care workers. **Table 1** shows the demographic and clinical characteristics of the patients.

**Table 1.** Patients characteristics. MV: mechanical ventilation; APACHE II: Acute Physiology and Chronic Health disease Classification System II; SOFA: Sequential Organ Failure Assessment; COPD: Chronic obstructive pulmonary disease.

|  |  | Non-MV | MV |  |
| --- | --- | --- | --- | --- |
|  | All patients<br>(n = 168) | Survivors<br>(n = 54) | Survivors<br>(n = 68) | Non-Survivors<br>(n = 46) |
| **Demographic data** |  |  |  |  |
| Male, no. (%) | 111 (66%) | 36 (66,6%) | 45 (66,2%) | 30 (65,2%) |
| Age, median (IQR), y | 67 (58-75) | 61 (54 -73) | 65 (59-73) | 71 (65-78) |
| Distribution, no. (%) |  |  |  |  |
| ≤ 39 years | 8 (4,7%) | 5 (9,2%) | 2 (2,9%) | 1 (2,1%) |
| 40-59 years | 44 (26,1%) | 19 (35,1%) | 17 (25%) | 8 (17,3%) |
| 60-79 years | 96 (57,1%) | 27 (50%) | 43 (63,2%) | 26 (56,5%) |
| ≥80 years | 20 (11,9%) | 3 (5,5%) | 6 (8,8%) | 11 (23,9%) |
| **Scores, median (IQR)** |  |  |  |  |
| APACHE II score | 10 (6-14) | 5 (3-10) | 11 (9-15) | 13 (10-17) |
| SOFA score day 1 | 2 (2-5) | 1 (1-2) | 3 (2-5) | 4 (2-5) |
| SOFA score day 3 | 2 (1-5) | 1 (1-2) | 3 (2-6) | 3 (2-6) |
| SOFA score day 5 | 3 (1-4) | 0 (0-1) | 3 (2-4) | 3 (2-4) |
| Charlson score | 4 (2-5) | 3 (1-5) | 4 (2-5) | 5 (4-7) |
| **Comorbidities, no. (%)** |  |  |  |  |
| Hypertension | 88 (52,4%) | 26 (48,1%) | 34 (50%) | 28 (60%) |
| Angiotensin-converting-enzyme inhibitors use | 31 (18,4%) | 11 (20,4%) | 14 (20,6%) | 6 (13%) |
| Angiotensin II receptor blocker use | 24 (14,3%) | 6 (11,1%) | 7 (10,3%) | 11 (23,9%) |
| Obesity | 70 (41,6%) | 25 (46,2%) | 29 (42,6%) | 16 (34,8%) |
| Diabetes | 31 (18,5%) | 12 (22,2%) | 10 (14,7%) | 9 (19,6%) |
| Immunosuppression | 26 (15,5%) | 7 (13%) | 6 (8,8%) | 13 (28,3%) |
| Active oncological disease without chemotherapy | 14 (8,3%) | 6 (11,3%) | 3 (4,5%) | 5 (10,9%) |
| Chronic use of corticosteroids | 11 (6,5%) | 2 (3,7%) | 4 (5,9%) | 5 (10,9%) |
| Active oncological disease with chemotherapy) | 9 (5,4%) | 2 (3,7%) | 1 (1,5%) | 6 (13%) |
| Solid organ transplant | 9 (5,4%) | 2 (3,7%) | 2 (2,9%) | 5 (10,9%) |
| Coronary heart disease | 24 (14,3%) | 7 (13%) | 10 (14,7%) | 7 (15,2%) |
| Chronic kidney disease | 20 (11,9%) | 4 (7,4%) | 8 (11,8%) | 8 (17,4%) |
| Active smoking | 18 (10,7%) | 5 (9,3%) | 8 (11,8%) | 5 (10,9%) |
| COPD | 10 (6%) | 3 (5,6%) | 2 (2,9%) | 5 (10,9%) |
| Asthma | 9 (5,4%) | 3 (5,6%) | 5 (7,4%) | 1 (2,2%) |
| Congestive heart failure | 5 (3%) | 2 (3,8%) | 2 (2,9%) | 1 (2,2%) |
| **Epidemiological link, no. (%)** |  |  |  |  |
| Community transmission | 140 (83,3%) | 44 (81,4%) | 58 (85,2%) | 38 (82,6%) |
| In-hospital transmission | 24 (14,3%) | 8 (14,8%) | 8 (11,7%) | 8 (17,4%) |
| Imported cases | 4 (2,4%) | 2 (3,7%) | 2 (3,7%) | 0 |
| **ICU admission criteria, no. (%)** |  |  |  |  |
| Clinical monitoring | 100 (59,5%) | 54 (100%) | 31 (46,3%) | 15 (32,6%) |
| Respiratory support | 68 (40,5%) | 0 | 37 (45,6%) | 31 (67,4%) |

The median (IQR) time between symptoms onset and hospital admission was 4 days (2-7). Fever was referred by 91,1% (153) of patients followed by shortness of breath (52.4% [88] patients). The main ICU admission criteria were clinical monitoring (59.5% of patients [100]), and 40.5% (68) of patients needed MV at ICU arrival. The median time up to ICU admission was 7 days (4-9) since symptoms onset.

### Radiologic and laboratory findings

All patients had chest x-rays at ICU admission, 98.2% (165) revealed abnormal results. The most common pattern on chest x-rays was bilateral patchy shadowing in 72% (121) with a median (IQR) RALE score of 7 (4-7). The presence of pleural effusion was infrequent (9.5% [16] of chest x-rays).

On ICU admission, lymphocytopenia was present in 73.2% (123) of patients, thrombocytopenia in 19.6% (33), and leukopenia in 6.5% (11).

Among MV survivors and MV non-survivors, no difference was found in total leukocyte count (median [IQR], 7292 mm^3^ [5867-9847] in MV survivors vs. 9004 mm^3^ [6455-12819] in MV non-survivors; *p*=0.1313) nor absolute neutrophil count (median [IQR], 6034 mm^3^ [4319-7827] in MV survivors vs. 7609 mm^3^ [5238-10980] in MV non-survivors; *p*=0.3366).

Regarding lymphocytes, absolute cell count was significantly higher among non-MV patients than MV patients (median [IQR], 867 mm^3^ [571-1148] vs. 668 mm^3^ [461-1005]; *p*=0.0042), and also, a significantly higher count was registered in MV survivors than MV non-survivors (499 mm^3^ in non-MV [352-862]; *p*=0.0294).

Neutrophil to lymphocyte ratio (NLR) was higher among non-MV patients than MV survivors in ventilated vs. non-ventilated patients (median [IQR], 6,9 [4,5-11,8] vs. 11,39 [5,5-24,2]; *p*=0.0059) but non difference was found between MV survivors vs. MV non-survivors (median [IQR], 11,2 [6,3-22,4] vs. 12,9 [4,9-25,3]; *p*=0.9913).

The majority of patients had elevated levels of inflammatory biomarkers like CRP, ferritin and IL-6. ProBNP was higher among ventilated patients vs. non-ventilated (median [IQR], 520 pg/ml [199-1189] vs. 172 [49-526]; *p*=0,0004). Also, among overall, non-surviving patients had more prominent laboratory abnormalities including proBNP and d-dimer than survivors (median [IQR], 690 pg/ml [362-2062] vs. 435 pg/ml [128-939]; *p*=0.0106 and 1158 ng/ml [826-1817] vs. ng/ml 818 [618-1305]; *p*=0,0097). **Table 2** shows the radiologic and laboratory findings at ICU admission. **Figure 1** shows laboratory results among non-MV, patients, MV survivors and MV non-survivors.

**Table 2.** Radiologic and laboratory findings at ICU admission. MV: mechanical ventilation; RALE: Radiographic Assessment of Lung Edema; ALP: Alkaline phosphatase; AST: Aspartate aminotransferase; ALT: Alanine aminotransferase; pro-BNP: Pro b-type natriuretic peptide; hs-cTnT: High-sensitivity cardiac troponin. ^a^Data regarding Procalcitonin were missing for 40 patients (24%). ^b^Data regarding pro-BNP were missing for 26 patients (15%). ^c^Data regarding hs-cTnT were missing for 74 patients (44%). ^d^Data regarding D-Dimer were missing for 41 patients (24%). ^e^Data regarding C-reactive protein were missing for 80 patients (48%). ^f^Data regarding Ferritin were missing for 77 patients (46%). ^g^Data regarding Lactate dehydrogenase were missing for 96 patients (57%). ^h^Data regarding Interleukin 6 were missing for 128 patients (76%).

|  |  | Non-MV | MV |  |
| --- | --- | --- | --- | --- |
|  | All patients<br>(n = 168) | Survivors<br>(n = 54) | Survivors<br>(n = 68) | Non-survivors<br>(n = 46) |
| **Radiologic findings** |  |  |  |  |
| RALE score | 7 (4-7) | 6 (3-7) | 7 (4-7) | 7 (5-7) |
| Bilateral compromise, no. (%) | 121 (78%) | 35 (64,8%) | 49 (72,1%) | 37 (80,4%) |
| Pleural effusion, no. (%) | 16 (9,5%) | 0 | 9 (13,2%) | 7 (15,2%) |
| **Laboratory findings, median (IQR)** |  |  |  |  |
| Hematocrit, % | 38 (34-42) | 40 (36-42) | 37 (34-41) | 38 (34-41) |
| Leukocytes, per mm3 | 8001 (5844-11713) | 7292 (5867-9847) | 9004 (6455-12819) | 7733 (5095-12688) |
| Neutrophils, per mm3 | 6560 (4464-10386) | 6034 (4319-7827) | 7609 (5238-10980) | 6302 (3901-11477) |
| Lymphocytes, per mm3 | 695 (456-1010) | 867 (571-1148) | 668 (461-1005) | 499 (352-862) |
| Neutrophils to Lymphocytes ratio | 9,5 (5,1-19,6) | 6,9 (4,5-11,8) | 11,2 (6,3-22,4) | 12,9 (4,9-25,3) |
| Platelets, per mm3 | 205000<br>(154000-267900) | 209200<br>(169625-266225) | 221100<br>(169750-280400) | 193200<br>(142800-231525) |
| Serum creatinine, mg/dL | 0,90 (0,71-1,18) | 0,83 (0,65-1,15) | 0,94 (0,71-1,12) | 1,05 (0,78-1,39) |
| Serum urea, mg/dL | 42 (31-57) | 37 (27-49) | 43 (32-58) | 47 (37-64) |
| Sodium, mmol/L | 135 (133-138) | 135 (132-137) | 136 (134-138) | 135 (133-139) |
| Potassium, mmol/L | 3,9 (3,7-4,4) | 3,8 (3,6-4,3) | 3,9 (3,6-4,4) | 4,0 (3,7-4,5) |
| Chloride, mmol/L | 102 (98-105) | 101 (98-103) | 103 (100-106) | 102 (98-106) |
| Total bilirubin, mg/dL | 0,58 (0,42-0,72) | 0,60 (0,47-0,74) | 0,58 (0,39-0,66) | 0,58 (0,42-0,88) |
| ALP, UI/L | 65 (52-89) | 61 (48-87) | 65 (52-80) | 75 (52-128) |
| AST, UI/L | 35 (27-48) | 33 (23-47) | 33 (25-48) | 38 (26-56) |
| ALT, UI/L | 29 (17-52) | 33 (18-58) | 26 (17-49) | 29 (17-45) |
| Albumin, g/dL | 3,37 (3,06-3,6) | 3,46 (3,26-3,80) | 3,23 (3,00-3,48) | 3,30 (2,94-3,58) |
| Prothrombin time, s | 85 (74-94) | 86 (78-93) | 85 (76-95) | 79 (64-94) |
| Procalcitonin, ng/mL<sup>a</sup> | 0,24 (0,09-0,66) | 0,17 (0,07-0,33) | 0,31 (0,09-0,61) | 0,36 (0,14-1,87) |
| pro-BNP, pg/mL<sup>b</sup> | 435 (118-1045) | 172 (49-526) | 435 (128-939) | 690 (362-2062) |
| hs-cTnT, pg/mL<sup>c</sup> | 14 (8-32) | 10 (7-16) | 13 (8-25) | 31 (14-92) |
| D-Dimer, ng/mL<sup>d</sup> | 945 (669-1529) | 740 (608-1300) | 947 (658-1344) | 1158 (826-1817) |
| C-reactive protein, mg/L<sup>e</sup> | 93 (51-151) | 82 (51-105) | 109 (59-140) | 99 (49-177) |
| Ferritin, g/mL<sup>f</sup> | 723 (384-1249) | 632 (267-1356) | 741 (409-1244) | 668 (422-1024) |
| Lactate dehydrogenase, UI/L<sup>g</sup> | 300 (256-355) | 264 (209-320) | 321 (272-379) | 313 (263-418) |
| Interleukin 6, pg/mL<sup>h</sup> | 55 (13-114) | 55 (11-104) | 48 (18-137) | 110 (14-271) |

**Figure 1.**
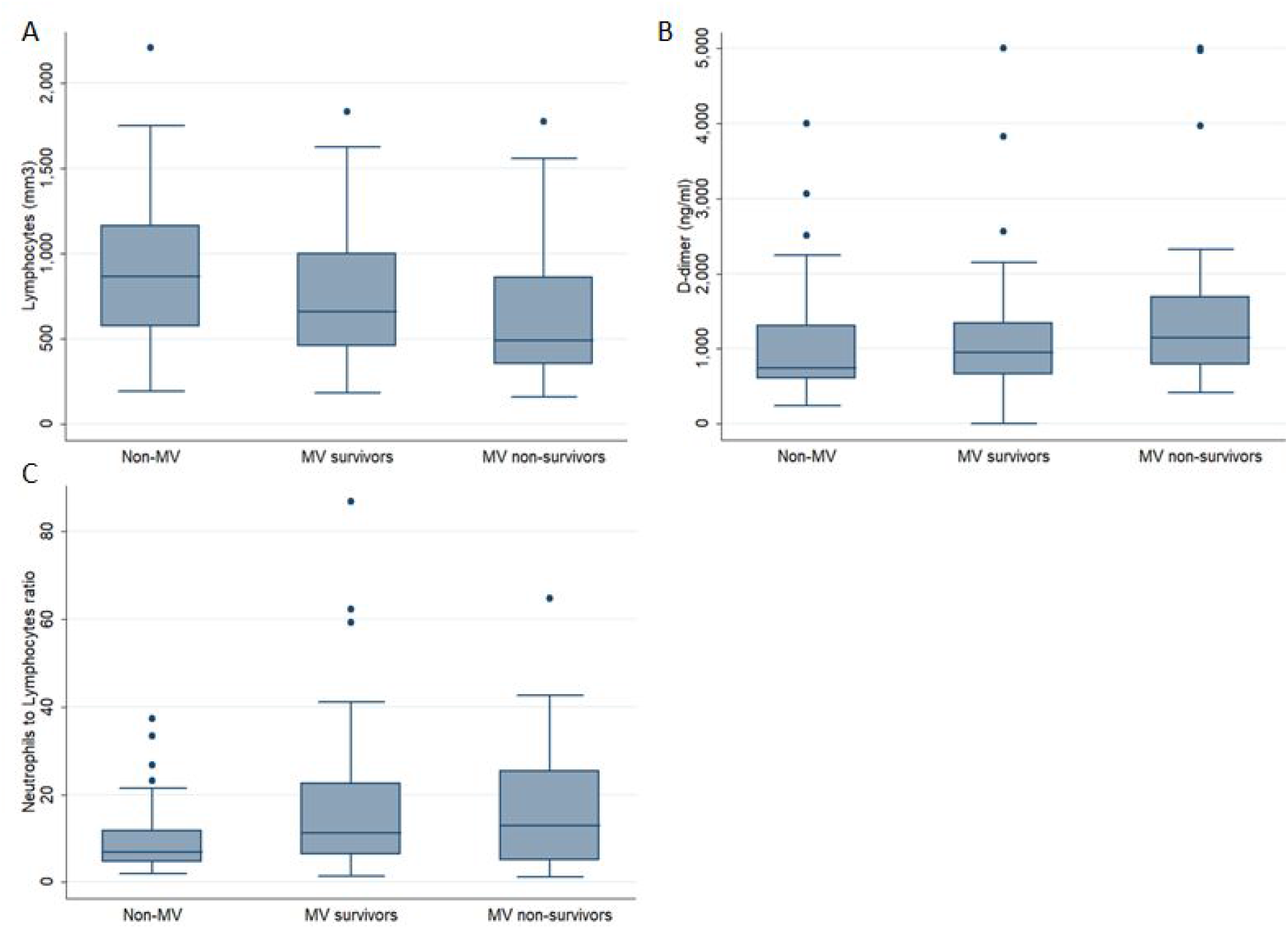
Laboratory results among non-mechanically ventilated (Non-MV) patients, MV survivors and MV non-survivors of, **A:** lymphocytes (per mm^3^); **B:** D-dimer (ng/ml); **C:** Neutrophils to Lymphocytes ratio

### Mechanical ventilation

A total of 114 patients (67.9%) required endotracheal intubation and invasive MV. No patient was treated with noninvasive ventilation. On the first day of MV, the median positive end-expiratory pressure (PEEP) was 10 (8-11) cm H_2_O. PEEP levels as high as 16 cm H_2_O were applied. Among a total of 114 patients, 75 (65.7%) required a fraction of inspired oxygen (FIO2) of at least 50%, and 11 (9.4%) required 100% FIO2. The median PaO2/FIO2 ratio was 200 (IQR, 147-268). Also, lower PaO2/FIO2 ratios on the first day of MV were registered among non-survivors vs. survivors (median [IQR], 180 [120-214] vs. 216 [167-290]; *p*=0.0309). All MV patients fulfilled Berlin criteria for Acute respiratory distress syndrome (ARDS)^7^.

Regarding respiratory mechanics, the median plateau pressure (P_plat_) on the first day of MV was 22 (19-24) cm H_2_O, the median driving pressure (ΔP) was 11 (10-14) cm H_2_O and the median respiratory system compliance (Crs) was 36 (30-47) ml/cm H_2_O.

A majority of the mechanically ventilated patients (84 [73.7%]) required neuromuscular blockade due to severe hypoxemia to avoid patient-ventilator asynchronies. Also, prone position ventilation was applied to 41 patients (36%), and 36,1% of them (15 patients) required more than one prone positioning session. 12 patients (10.5%) received inhaled nitric oxide (iNO) and 6 (5.7%) were connected to veno-venous extracorporeal membrane oxygenation (V-V ECMO) because of refractory hypoxemia.

The time elapse of MV was 16 days (19-30) among overall ventilated patients and 8 days (6-14) among those who were successfully extubated (26 [22.8%]). 32 patients (28.1%) underwent percutaneous tracheostomy due to prolonged weaning. The median time of MV until tracheostomy was 20 days (15-27). 10 patients (8.7%) remained under MV at data cut off. **Table 3** shows respiratory mechanics, oxygenation parameters and adjunctive therapies applied.

**Table 3.**
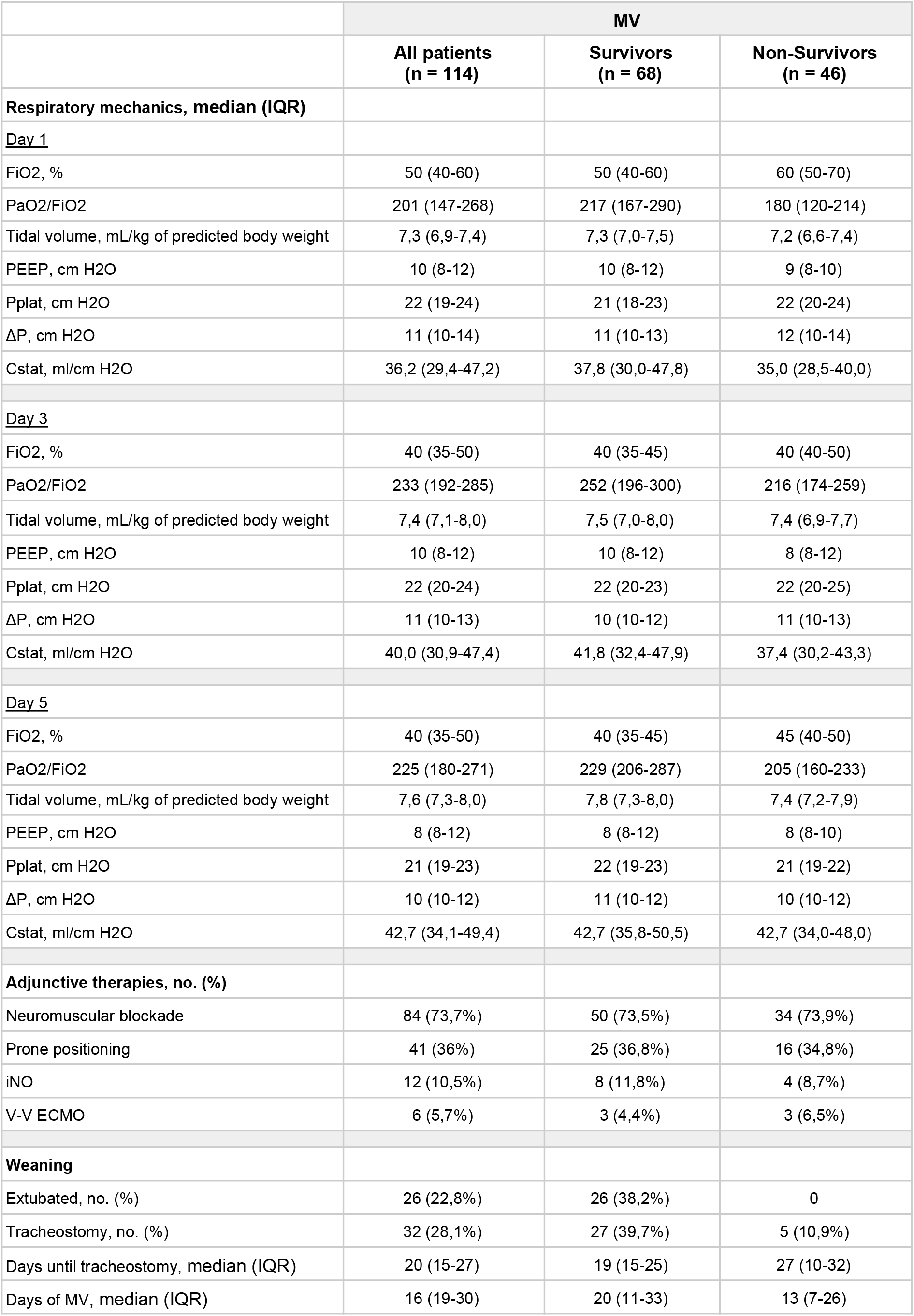
Respiratory mechanics, oxygenation parameters and adjunctive therapies applied. MV: mechanical ventilation; FiO2: Fraction of inspired oxygen; PaO2: Partial pressure of oxygen; PEEP: Positive end-expiratory pressure; Pplat: Plateau pressure; ΔP: Driving pressure; Crs: respiratory system compliance; iNO: Inhaled nitric oxide; V-V ECMO: veno-venous extracorporeal membrane oxygenation.

|  | MV |  |  |
| --- | --- | --- | --- |
|  | All patients<br>(n = 114) | Survivors<br>(n = 68) | Non-Survivors<br>(n = 46) |
| **Respiratory mechanics, median (IQR)** |  |  |  |
| <u>Day 1</u> |  |  |  |
| FiO2, % | 50 (40-60) | 50 (40-60) | 60 (50-70) |
| PaO2/FiO2 | 201 (147-268) | 217 (167-290) | 180 (120-214) |
| Tidal volume, mL/kg of predicted body weight | 7,3 (6,9-7,4) | 7,3 (7,0-7,5) | 7,2 (6,6-7,4) |
| PEEP, cm H2O | 10 (8-12) | 10 (8-12) | 9 (8-10) |
| Pplat, cm H2O | 22 (19-24) | 21 (18-23) | 22 (20-24) |
| ΔP, cm H2O | 11 (10-14) | 11 (10-13) | 12 (10-14) |
| Cstat, ml/cm H2O | 36,2 (29,4-47,2) | 37,8 (30,0-47,8) | 35,0 (28,5-40,0) |
| <u>Day 3</u> |  |  |  |
| FiO2, % | 40 (35-50) | 40 (35-45) | 40 (40-50) |
| PaO2/FiO2 | 233 (192-285) | 252 (196-300) | 216 (174-259) |
| Tidal volume, mL/kg of predicted body weight | 7,4 (7,1-8,0) | 7,5 (7,0-8,0) | 7,4 (6,9-7,7) |
| PEEP, cm H2O | 10 (8-12) | 10 (8-12) | 8 (8-12) |
| Pplat, cm H2O | 22 (20-24) | 22 (20-23) | 22 (20-25) |
| ΔP, cm H2O | 11 (10-13) | 10 (10-12) | 11 (10-13) |
| Cstat, ml/cm H2O | 40,0 (30,9-47,4) | 41,8 (32,4-47,9) | 37,4 (30,2-43,3) |
| <u>Day 5</u> |  |  |  |
| FiO2, % | 40 (35-50) | 40 (35-45) | 45 (40-50) |
| PaO2/FiO2 | 225 (180-271) | 229 (206-287) | 205 (160-233) |
| Tidal volume, mL/kg of predicted body weight | 7,6 (7,3-8,0) | 7,8 (7,3-8,0) | 7,4 (7,2-7,9) |
| PEEP, cm H2O | 8 (8-12) | 8 (8-12) | 8 (8-10) |
| Pplat, cm H2O | 21 (19-23) | 22 (19-23) | 21 (19-22) |
| ΔP, cm H2O | 10 (10-12) | 11 (10-12) | 10 (10-12) |
| Cstat, ml/cm H2O | 42,7 (34,1-49,4) | 42,7 (35,8-50,5) | 42,7 (34,0-48,0) |
| **Adjunctive therapies, no. (%)** |  |  |  |
| Neuromuscular blockade | 84 (73,7%) | 50 (73,5%) | 34 (73,9%) |
| Prone positioning | 41 (36%) | 25 (36,8%) | 16 (34,8%) |
| iNO | 12 (10,5%) | 8 (11,8%) | 4 (8,7%) |
| V-V ECMO | 6 (5,7%) | 3 (4,4%) | 3 (6,5%) |
| **Weaning** |  |  |  |
| Extubated, no. (%) | 26 (22,8%) | 26 (38,2%) | 0 |
| Tracheostomy, no. (%) | 32 (28,1%) | 27 (39,7%) | 5 (10,9%) |
| Days until tracheostomy, median (IQR) | 20 (15-27) | 19 (15-25) | 27 (10-32) |
| Days of MV, median (IQR) | 16 (19-30) | 20 (11-33) | 13 (7-26) |

### Treatment and complications

Most of the patients received dexamethasone (on RECOVERY-trial dose^9^) and empirical intravenous antibiotic therapy (143 [85,1%] and 133 [79,2%], respectively). 90 patients (53,6%) were included in a 2:1 blinded-protocol of convalescent plasma against placebo. Other treatments like ritonavir/lopinavir (23 [13.7%]) and tocilizumab (3 [1.8%]) were less frequently applied.

A common ICU related complication was delirium that was identified in 121 patients (72%) overall. Delirium was less present among non-ventilated patients (23 [42.5%]) in comparison with ventilated patients that survived (61 [89,7%]) and those who died (37 [80.4%]). Other complications registered were catheter-related sepsis (23 [13.7%]), pressure ulcers (17 [20.1%]), urosepsis (10 [6%]) and ventilator-associated pneumonia (11 [6.5%])

Among overall patients, the median duration of hospitalization was 21 days (14-32) and the median length of stay in ICU was 13 days (6-24). ICU length of stay among survivors was 5 days (3-8) for non-ventilated patients and 23 days (16-31) for those patients who required invasive MV. Overall ICU mortality of this series was 25% (42). **Table 4** shows treatments, ICU related complications, and clinical outcomes at data cutoff.

**Table 4.**
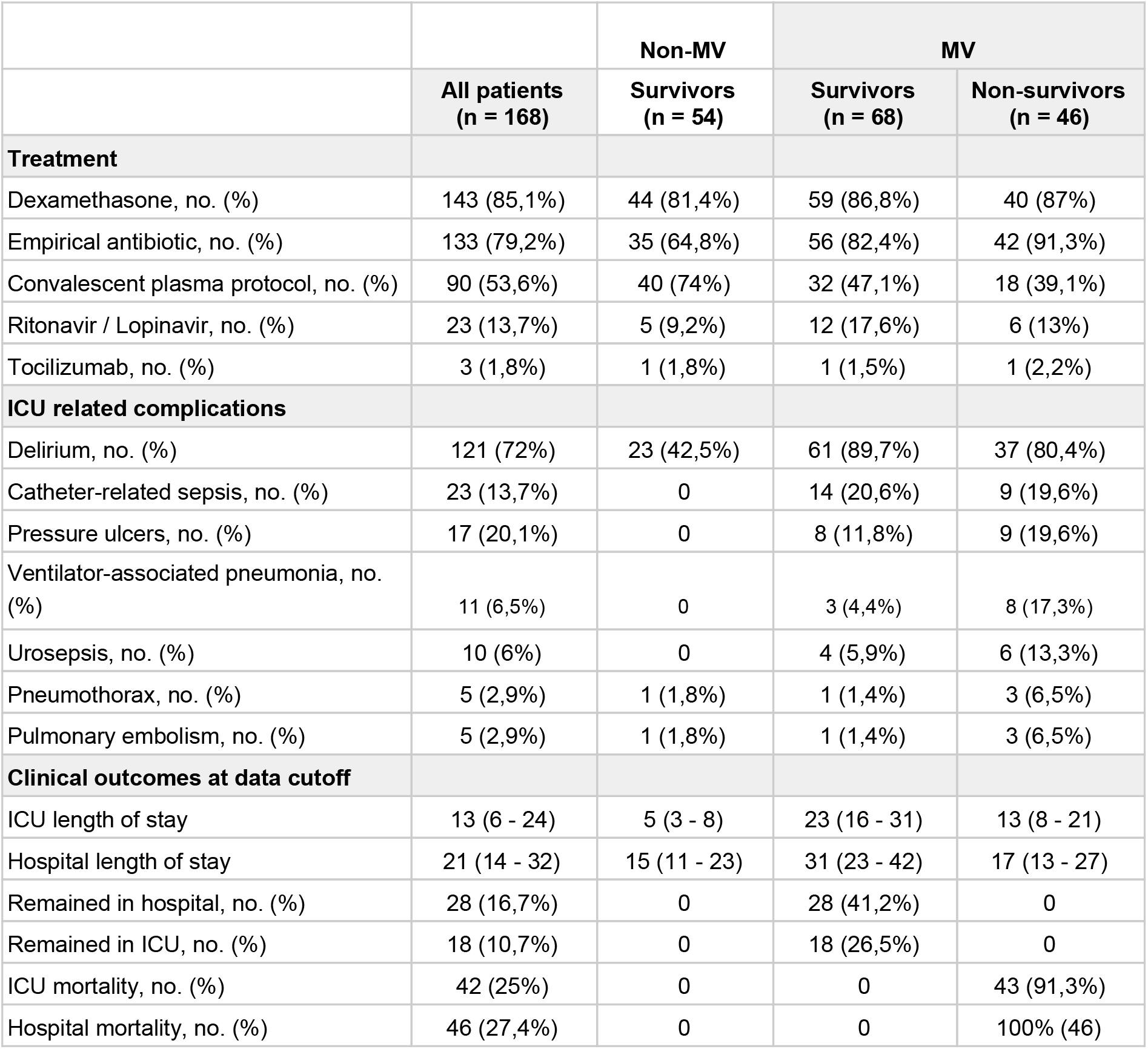
Treatment, ICU related complications and clinical outcomes. MV: mechanical ventilation; ICU: intensive care unit.

|  |  | Non-MV | MV |  |
| --- | --- | --- | --- | --- |
|  | All patients<br>(n = 168) | Survivors<br>(n = 54) | Survivors<br>(n = 68) | Non-survivors<br>(n = 46) |
| **Treatment** |  |  |  |  |
| Dexamethasone, no. (%) | 143 (85,1%) | 44 (81,4%) | 59 (86,8%) | 40 (87%) |
| Empirical antibiotic, no. (%) | 133 (79,2%) | 35 (64,8%) | 56 (82,4%) | 42 (91,3%) |
| Convalescent plasma protocol, no. (%) | 90 (53,6%) | 40 (74%) | 32 (47,1%) | 18 (39,1%) |
| Ritonavir / Lopinavir, no. (%) | 23 (13,7%) | 5 (9,2%) | 12 (17,6%) | 6 (13%) |
| Tocilizumab, no. (%) | 3 (1,8%) | 1 (1,8%) | 1 (1,5%) | 1 (2,2%) |
| **ICU related complications** |  |  |  |  |
| Delirium, no. (%) | 121 (72%) | 23 (42,5%) | 61 (89,7%) | 37 (80,4%) |
| Catheter-related sepsis, no. (%) | 23 (13,7%) | 0 | 14 (20,6%) | 9 (19,6%) |
| Pressure ulcers, no. (%) | 17 (20,1%) | 0 | 8 (11,8%) | 9 (19,6%) |
| Ventilator-associated pneumonia, no. (%) | 11 (6,5%) | 0 | 3 (4,4%) | 8 (17,3%) |
| Urosepsis, no. (%) | 10 (6%) | 0 | 4 (5,9%) | 6 (13,3%) |
| Pneumothorax, no. (%) | 5 (2,9%) | 1 (1,8%) | 1 (1,4%) | 3 (6,5%) |
| Pulmonary embolism, no. (%) | 5 (2,9%) | 1 (1,8%) | 1 (1,4%) | 3 (6,5%) |
| **Clinical outcomes at data cutoff** |  |  |  |  |
| ICU length of stay | 13 (6 - 24) | 5 (3 - 8) | 23 (16 - 31) | 13 (8 - 21) |
| Hospital length of stay | 21 (14 - 32) | 15 (11 - 23) | 31 (23 - 42) | 17 (13 - 27) |
| Remained in hospital, no. (%) | 28 (16,7%) | 0 | 28 (41,2%) | 0 |
| Remained in ICU, no. (%) | 18 (10,7%) | 0 | 18 (26,5%) | 0 |
| ICU mortality, no. (%) | 42 (25%) | 0 | 0 | 43 (91,3%) |
| Hospital mortality, no. (%) | 46 (27,4%) | 0 | 0 | 100% (46) |

## DISCUSSION

The population in this study consisted mostly of men (66%) with a median age of 67 (58-75) years old, which is substantially high compared to the median age of all the positive Argentinian cases of COVID-19 (38 years old)^10^. The previous suggests that gender and age are risk factors for admission to the ICU, as previously reported^11^. Moreover, in this cohort, most of the patients (79.7%) had at least one comorbidity, with a large proportion of patients with hypertension (52.4%) and obesity (41,6%).

In this case series, the majority of patients were admitted to the ICU because of acute hypoxemic respiratory failure that required clinical monitoring. An important proportion of patients needed respiratory support and endotracheal intubation at ICU arrival. Endotracheal intubation and invasive MV were needed in 67.9% of the patients, whereas only 32.1% could be managed with oxygen delivery by a non-rebreather mask. Setting primary focus on health-care personnel security, no patient received non-invasive MV (eg. continuous positive airway pressure, non-invasive positive pressure ventilation, or high flow nasal cannula) due to the risk of aerosol dispersion^12^. There is still controversy regarding the efficacy of non-invasive MV devices to avoid endotracheal intubation^13^ but the need for invasive MV in this patient population under study was similar to other ICUs where non-invasive MV was applied: 65% (São Paulo, Brazil)^14^, 71% (Washington State, United States)^15^, 71% (Wuhan, China)^16^, 80.8% (Detroit, United States)^17^, 88% (Lombardy region, Italy)^18^, 96.7% (Mexico City, Mexico)^19^.

Among ventilated patients, 44.7% (51) presented paO2/FiO2 lower than 200 on the first day of MV, although pulmonary mechanics were almost normal (median P_plat_ 22 cm H_2_O [19-24] and median ΔP 11 cm H_2_O [10-14]). Nevertheless, 79.8% (91 patients) had low Crs (<50 ml/cm H_2_O) with a median Crs of 36 ml/cm H_2_O (30-47) despite of lung-protective ventilation (tidal volume ≤ 8 ml/kg predicted body-weight). These findings are in accordance with information that reported that COVID-19 ARDS respiratory features are similar to non-COVID-19 ARDS^20^. Additionally, the values of Crs, Pplat, and ΔP were very similar to previously published cohorts of non-COVID-19 ARDS patients^21^.

Due to refractory hypoxemia, 6 patients (5.7%) were connected to V-V ECMO. Despite the middle-income country circumstances and the low volume of patients of the ECMO program, outcomes are comparable to international series. ECMO survival and survival to discharge was 66% and 50%, respectively^22,23,24^.

Since patients required MV for a long period of time, due to prolonged weaning, there were more that underwent percutaneous tracheostomy compared to those who were successfully extubated (32 patients [28.1%] and 26 patients [22.8%], respectively). At the beginning of the pandemic, several scientific societies warned about a possible increase in the request for tracheostomies related to SARS-CoV-2 infection^25,26,27,29^. The need for tracheostomies in critically ill COVID-19 patients ranges from 36% to 53%, according to the data found in the literature^28–30^.

Prioritizing the protection of health personnel, tracheostomy was considered possible only in patients who were expected to obtain a substantial benefit. Ideally, it was performed 21 days after intubation with a negative RT-PCR test^31^. Following that criteria and up to data cutoff, none of the surgeons or intensivists who performed tracheostomies developed symptoms or tested positive for COVID-19. Nevertheless, a longer time of MV prior to tracheostomy was observed in relation to a previous series from the same center in a non-pandemic scenario (median 20 days [15-27] and 9 days [6-12], respectively)^32^.

ICU mortality rate was slightly lower than a recent systematic review and meta-analysis of observational studies that informed a 41.6% (34.0-49.7) ICU mortality rate across international studies. This could be explained because, optimistically, countries in the later phase of the pandemic may be coping better with COVID-19^33^.

Although this study is considered limited due to the relatively small number of patients from a single center and could not be broadly applicable to other patients with critical illness, it definitely provides initial experience regarding characteristics of COVID-19 in patients with critical illness in South America.

## CONCLUSION

In Argentina in a high complexity hospital in the city of Buenos Aires, the study of critically ill patients infected with SARS-CoV-2, a majority of which were comorbid elder men that required MV in a large proportion, showed that global ICU mortality was maintained at 25%.

## Data Availability

All data is available.

## Acknowledgements

The research team is especially grateful to Jorge Luis Aliaga and Maria de los Ángeles Magaz for their valuable collaboration in this work.

## REFERENCES

1. WHO Director-General’s opening remarks at the media briefing on COVID-19-11 March 2020, https://www.who.int/dg/speeches/detail/who-director-general-s-opening-remarks-at-the-media-briefing-on-covid-19---11-march-2020 (accessed 27 September 2020).

2. Gemelli NA. Management of COVID-19 Outbreak in Argentina: The Beginning. Disaster Med Public Health Prep 2020; 1–3.

3. Septiembre de 2020, https://www.argentina.gob.ar/coronavirus/informes-diarios/reportes/septiembre2020 (2020, accessed 27 September 2020).

4. Carboni Bisso I, Huespe I, Lockhart C, et al. [COVID-19 in the intensive care unit. Analysis of the experience during the first month of pandemic]. Medicina 2020; 80 Suppl 3: 25–30.

5. [World Health Organization], https://apps.who.int/iris/handle/10665/330854 (accessed 27 September 2020).

6. Jabaudon M, Audard J, Jaber S, et al. The Radiographic Assessment of Lung Edema (RALE) Score Is Associated with Survival and May Be Useful to Identify Focal and Non-Focal Lung Imaging Phenotypes in Patients with ARDS. A25. ARDS: NEW APPROACHES TO DIAGNOSIS AND TREATMENT. Epub ahead of print 2020. DOI: 10.1164/ajrccm-conference.2020.201.1_meetingabstracts.a1133.

7. ARDS Definition Task Force, Ranieri VM, Rubenfeld GD, et al. Acute respiratory distress syndrome: the Berlin Definition. JAMA 2012; 307: 2526–2533.

8. Khwaja A. KDIGO clinical practice guidelines for acute kidney injury. Nephron Clin Pract 2012; 120: c179–84.

9. Group TRC, The RECOVERY Collaborative Group. Dexamethasone in Hospitalized Patients with Covid-19 — Preliminary Report. New England Journal of Medicine. Epub ahead of print 2020. DOI: 10.1056/nejmoa2021436.

10. Ministerio de Salud de la Nación Argentina, https://www.argentina.gob.ar/sites/default/files/sala_29_09_.pdf (accessed 30 September 2020).

11. Williamson EJ, Walker AJ, Bhaskaran K, et al. Factors associated with COVID-19-related death using OpenSAFELY. Nature 2020; 584: 430–436.

12. Ferioli M, Cisternino C, Leo V, et al. Protecting healthcare workers from SARS-CoV-2 infection: practical indications. Eur Respir Rev; 29. Epub ahead of print 31 March 2020. DOI: 10.1183/16000617.0068-2020.

13. Agarwal A, Basmaji J, Muttalib F, et al. High-flow nasal cannula for acute hypoxemic respiratory failure in patients with COVID-19: systematic reviews of effectiveness and its risks of aerosolization, dispersion, and infection transmission. Can J Anaesth 2020; 67: 1217–1248.

14. Teich VD, Klajner S, Almeida FAS de, et al. Epidemiologic and clinical features of patients with COVID-19 in Brazil. Einstein 2020; 18: eAO6022.

15. Arentz M, Yim E, Klaff L, et al. Characteristics and Outcomes of 21 Critically Ill Patients With COVID-19 in Washington State. JAMA 2020; 323: 1612–1614.

16. Basu A, Chakraborty S. Faculty Opinions recommendation of Clinical course and outcomes of critically ill patients with SARS-CoV-2 pneumonia in Wuhan, China: a single-centered, retrospective, observational study. Faculty Opinions – Post-Publication Peer Review of the Biomedical Literature. Epub ahead of print 2020. DOI: 10.3410/f.737422643.793575066.

17. Suleyman G, Fadel RA, Malette KM, et al. Clinical Characteristics and Morbidity Associated With Coronavirus Disease 2019 in a Series of Patients in Metropolitan Detroit. JAMA Network Open 2020; 3: e2012270.

18. Grasselli G, Zangrillo A, Zanella A, et al. Baseline Characteristics and Outcomes of 1591 Patients Infected With SARS-CoV-2 Admitted to ICUs of the Lombardy Region, Italy. JAMA 2020; 323: 1574–1581.

19. Mendez-Probst CE, Velázquez-Fernández D, Castillejos-Molina RA, et al. Clinical and Epidemiological Characteristics of Patients Diagnosed with COVID-19 in a Tertiary Care Center in Mexico City: A Prospective Cohort Study. Revista de investigación Clínica; 72. Epub ahead of print 2020. DOI: 10.24875/ric.20000301.

20. Ferrando C, Suarez-Sipmann F, Mellado-Artigas R, et al. Clinical features, ventilatory management, and outcome of ARDS caused by COVID-19 are similar to other causes of ARDS. Intensive Care Med. Epub ahead of print 29 July 2020. DOI: 10.1007/s00134-020-06192-2.

21. Bellani G, Laffey JG, Pham T, et al. Epidemiology, Patterns of Care, and Mortality for Patients With Acute Respiratory Distress Syndrome in Intensive Care Units in 50 Countries. JAMA 2016; 315: 788–800.

22. Laffey JG, Madotto F, Bellani G, et al. Geo-economic variations in epidemiology, patterns of care, and outcomes in patients with acute respiratory distress syndrome: insights from the LUNG SAFE prospective cohort study. Lancet Respir Med 2017; 5: 627–638.

23. Riera J, Argudo E, Martínez-Martínez M, et al. Extracorporeal Membrane Oxygenation Retrieval in Coronavirus Disease 2019: A Case-Series of 19 Patients Supported at a High-Volume Extracorporeal Membrane Oxygenation Center. Critical Care Explorations 2020; 2: e0228.

24. Schmidt M, Hajage D, Lebreton G, et al. Extracorporeal membrane oxygenation for severe acute respiratory distress syndrome associated with COVID-19: a retrospective cohort study. Lancet Respir Med. Epub ahead of print 13 August 2020. DOI: 10.1016/S2213-2600(20)30328-3.

25. Tracheostomy guidance during the COVID-19 Pandemic, https://www.entuk.org/tracheostomy-guidance-during-covid-19-pandemic (accessed 1 October 2020).

26. Ralli M, Greco A, de Vincentiis M. The Effects of the COVID-19/SARS-CoV-2 Pandemic Outbreak on Otolaryngology Activity in Italy. Ear Nose Throat J 2020; 145561320923893.

27. Piazza C, Filauro M, Dikkers FG, et al. Long-term intubation and high rate of tracheostomy in COVID-19 patients might determine an unprecedented increase of airway stenoses: a call to action from the European Laryngological Society. European Archives of Oto-Rhino-Laryngology. Epub ahead of print 2020. DOI: 10.1007/s00405-020-06112-6.

28. Angel L, Kon ZN, Chang SH, et al. Novel Percutaneous Tracheostomy for Critically Ill Patients With COVID-19. Ann Thorac Surg 2020; 110: 1006–1011.

29. Picetti E, Fornaciari A, Taccone FS, et al. Safety of bedside surgical tracheostomy during COVID-19 pandemic: A retrospective observational study. PLoS One 2020; 15: e0240014.

30. Volo T, Stritoni P, Battel I, et al. Elective tracheostomy during COVID-19 outbreak: to whom, when, how? Early experience from Venice, Italy. Eur Arch Otorhinolaryngol. Epub ahead of print 12 July 2020. DOI: 10.1007/s00405-020-06190-6.

31. Smith D, Montagne J, Raices M, et al. Tracheostomy in the intensive care unit: Guidelines during COVID-19 worldwide pandemic. Am J Otolaryngol 2020; 41: 102578.

32. Carboni Bisso I, Huespe I, Schverdfinger S, et al. Traqueostomía percutánea guiada por broncoscopía: experiencia en 235 procedimientos. Revista de la Facultad de Ciencias Médicas de Córdoba 2020; 77: 187–190.

33. Armstrong RA, Kane AD, Cook TM. Outcomes from intensive care in patients with COVID-19: a systematic review and meta-analysis of observational studies. Anaesthesia 2020; 75: 1340–1349.

